# A Syndrome of Joint Hypermobility, Autonomic Dysfunction, Gastrointestinal Dysfunction and Autoimmune markers (JAG-A): Clinical Associations and Response to Intravenous Immunoglobulin Therapy

**DOI:** 10.1101/2023.10.01.23296388

**Authors:** Pankaj J. Pasricha, Megan McKnight, Luisa Villatoro, Guillermo Barahona, Jeffrey Brinker, Ken Hui, Michael Polydefkis, Robert Burns, Zsuzsanna H. McMahan, Neda Gould, Brent Goodman, Joseph Hentz, Glenn Treisman

## Abstract

**Background and aims:** We examined autoimmunity markers (AIMs) in patients with unexplained gastrointestinal symptoms, their relationship to joint hypermobility/hypermobility spectrum disorder (JH/HSD) and the response to intravenous immunoglobulin (IVIG).

**Methods:** The study comprised of three cohorts, consisting of adolescent or adult patients with gastrointestinal symptoms affecting more than one region of the gut who underwent laboratory tests, whole gut transit studies, and autonomic testing. AIM positive patients were defined based on a diagnosis of known rheumatic disease with one positive seromarker of autoimmunity or at least two positive seromarkers. The three cohorts were (a) Retrospective (n = 300); (b) Prospective validation cohort (n =133); and (c) Patients with AIM (n=32) prospectively treated with IVIG and followed with standardized questionnaires.

**Results:** AIMs were found in 39% of the retrospective cohort, of which the majority had a known rheumatic disorder. In the prospective cohort AIMs were noted in 35% overall but the rate was much higher in patients with JH/HSD (49% versus 21%, p=0.001). Significantly more patients with AIMs had elevations of C-reactive protein and erythrocyte sedimentation rate along with trends in tilt table test and HLADQ8 positivity. IVIG treatment was associated with a significantly greater overall treatment effect than controls and robust improvement over baseline in pain, gastrointestinal and autonomic symptoms.

**Conclusions:** Autoimmune markers and autonomic dysfunction are common in patients with unexplained gastrointestinal dysmotility, especially in those with joint hypermobility. IVIG treatment was associated with symptomatic improvement in both gastrointestinal and autonomic symptoms. These results need to be corroborated by randomized clinical trials of immunomodulators but suggest that an autoimmune etiology may be important to diagnose in such patients. Clinicaltrials.gov, NCT04859829

## Introduction

An autoimmune etiology or association is often present in disorders affecting the peripheral motor, sensory and autonomic nervous systems. Patients presenting primarily with these syndromes, are commonly also noted to have gastrointestinal dysmotility.^1, 2^ Gastrointestinal and autonomic dysfunction has also been seen in greater frequency in patients with joint hypermobility (JH/HSD).^3, 4^ Despite these known associations, patients whose primary complaints are gastrointestinal in nature are seldom investigated in detail for co-morbid autonomic dysfunction, joint hypermobility or an underlying autoimmune disorder. In the absence of known genetic disease (such as mitochondrial disorders) or diabetes, these patients are instead thought to be suffering from various so-called “functional” conditions labeled as irritable bowel syndrome, functional dyspepsia, idiopathic constipation and others or motility disorders such as idiopathic gastroparesis.

Auto-antibodies have been described previously in patients with unexplained GI dysmotility or so-called “functional” symptoms in a variety of settings including small series of patients with constipation and gastroparesis.^5,6^ However, antibodies that have been shown to be pathogenic are very few^7, 8^ ^5, 9, 10^, although numerous case series of patients with GI dysmotility have been described where surgically obtained specimens show an inflammatory (lymphocytes or eosinophils) infiltrate around damaged enteric neurons or muscle, with or without circulating antibodies.^11, 12^ Uncovering an immunological basis for the symptoms in these patients is important because of the lack of satisfactory treatment options particularly those that may be disease-modifying. Our principal aim was therefore to examine markers of autoimmunity and their association with JH/HSD and dysautonomia in a large patient population with refractory gastrointestinal symptoms without an obvious etiology. To further test the role of putative autoimmunity in the pathogenesis of these symptoms, we also prospectively treated a cohort of these patients with empirical immunomodulation using intravenous immunoglobulin (IVIG) in a small proof-of-concept trial.

## Methods

### Patients and Treatments

#### Definition of “autoimmune-marker (AIM) positivity”

This was defined as a known diagnosis of a canonical autoimmune rheumatic disease with at least one circulating seromarker associated with autoimmunity, or in patients not meeting classical criteria for a rheumatic disease, having two or more such seromarkers.

### Retrospective Cohort

A chart review was initially conducted on a consecutive cohort of adolescent or adult patients presenting to a single experienced gastroenterologist at a tertiary referral center seen during a period of approximately 2 years. The primary inclusion criterion was the presence of unexplained gastrointestinal symptoms (including pain) reflecting more than one region of the gut (esophagus, stomach, small intestine, or colon). Patients with single organ symptomatology (e.g., dysphagia due to achalasia or reflux symptoms only), inflammatory bowel disease, chronic pancreatitis, or other likely etiological factors (such as uncontrolled diabetes, adrenal insufficiency, malignancy, Parkinson’s Disease or other neurological disorder, uncontrolled thyroid disorder) were excluded. Patients routinely underwent serological testing for multiple commercially available antibodies and whole gastrointestinal transit by scintigraphy. In addition, patients with suspected dysautonomia underwent cardiovascular responses by provocative tilt table testing, and skin biopsies to assess for sudomotor and small fiber neuropathy. Details of these methods are provided in the Appendix.

### Prospective (validation) cohort

After an initial analysis of the prospective cohort, new patients presenting to the same clinic with the same inclusion/exclusion criteria as above were invited to participate in a patient registry with the aims of validating the findings as well as providing more granular clinical information through the use of standardized patient reported outcome questionnaires. These patients were seen by the same gastroenterologist, usually in a multi-disciplinary clinic also staffed by a psychiatrist/pain management specialist (GT), a clinical psychologist (NG), and rheumatologist (ZM). Subjects were administered the following questionnaires on a REDCAP platform prior to or at the time of their first clinic visit: (1) Patient Assessment of Upper Gastrointestinal Disorders Symptom Severity Index (PAGI-SYM)^13–15^ (2) PAGI-Quality of Life (PAGI-QOL)^16^ (3) Patient assessment of constipation (PAC-SYM)^17^ (4) Patient Assessment of Constipation-Quality of Life (PAC-QOL) questionnaire^18^ (5) Pelvic Floor Distress Inventory (PFDI)^19^ (6) Composite Autonomic Symptom Score (COMPASS-31)^20^ which was also used to define the presence or absence of dysautonomia, based on a cut-off of 32 which was the maximum score in a sample of thirty normal patients^21^ (7) 5-point questionnaire (5-PQ) score for joint hypermobility(JH)^22^ used to define patients with what is now preferred to be called Hypermobility Spectrum Disorder (HSD)^23^ (8) Eckardt symptom score for dysphagia^24^ (9) Short form McGill Pain Questionnaire (SF-MPQ)^25^ (10) Multiple Patient-Reported Outcomes Measurement Information System (PROMIS) measures^26^ and (11) Rome IV diagnostic questionnaire for adults^27^. Details of these questionnaires are provided in the Appendix.

### Treatment cohort

In a separate study, patients suspected to have autoimmune disease with unexplained and severe gastrointestinal complaints refractory to both standard and off-label symptomatic therapies were treated with IVIG by a single physician (PJP) and followed prospectively. IVIG was administered at a standard immunomodulatory dose (2 gm/kg body weight administered monthly). Clinical symptoms were assessed at baseline using many of the above standardized validated questionnaires and after every month and a 15-point overall treatment effectiveness (OTE) questionnaire.^28^ Patients who completed at least 3 monthly courses of IVIG were included in the final analysis.

### Statistical Methods

For the prospective cohort, symptom scores and demographics were compared between patients with and without autoimmunity, and between patients with and without joint hypermobility. Statistical significance was calculated by using the two-sample *t* test or Pearson chi-square test. Multiple comparisons were assessed by using the FDR (false discovery rate) method. Objective tests and demographics were compared between patients with and without autoimmunity in a pooled sample from the prospective and retrospective cohorts. Statistical significance was calculated by using Pearson chi-square test or two-sample *t* test. Multiple comparisons were assessed by using the FDR method. For the treatment cohort, the primary outcome measure was the Overall Treatment Effect score, which is a measure of change reported on a scale from -7 (worst) to 7 (best) points. The mean score was compared to zero by using the one-sample *t* test. Secondary outcome measures included other scale and subscale scores. Mean secondary scores after treatment were compared to those before treatment by using the paired *t* test. Statistical significance for the secondary measures was tested if the primary outcome was significant. The Hochberg method was then used to account for multiple comparisons among the 11 secondary scales. In addition, mean scores for patients with IVIG treatment were compared to those in the untreated group. Statistical significance was calculated by using the two-sample *t* test. The relationships between baseline characteristics and Overall Treatment Effect score were assessed by using the Pearson or point biserial correlation coefficient. In addition, baseline characteristics of patients with an Overall Treatment Effect score of at least “3” were compared to those with a score less than “3” and statistical significance was calculated by using the two-sample *t* test or Pearson chi-square test. Computations were performed using R software version 4.2.2.

### Human Subjects Approval

The entire study was approved by the Institutional Review Board of the Johns Hopkins University School of Medicine and the IVIG trial was registered with ClinicalTrials.gov (NCT04859829).

## Results

### Retrospective Cohort

Three hundred (300) consecutive patients were included in this analysis. As a group these patients were on average 40.5 (± 16.7) years old and 89% of them were female. The presenting complaint(s) in order of frequency were abdominal pain (89%), bloating/distention (88%), constipation (79%), nausea (78%), early satiety/fullness (69%), vomiting (50%), dysphagia (36%) and diarrhea (34%). Postural symptoms (dizziness, lightheadedness, syncope) were reported by 77% of patients and abnormal sweating by 53%. A history of migraine/recurrent headaches was present in 74%. One hundred and seventeen (117) patients (39%) were positive for AIM, of which 85 (73%) had a history of a known autoimmune diagnosis (Table 1), along with at least one autoantibody marker.

**Table 1:**
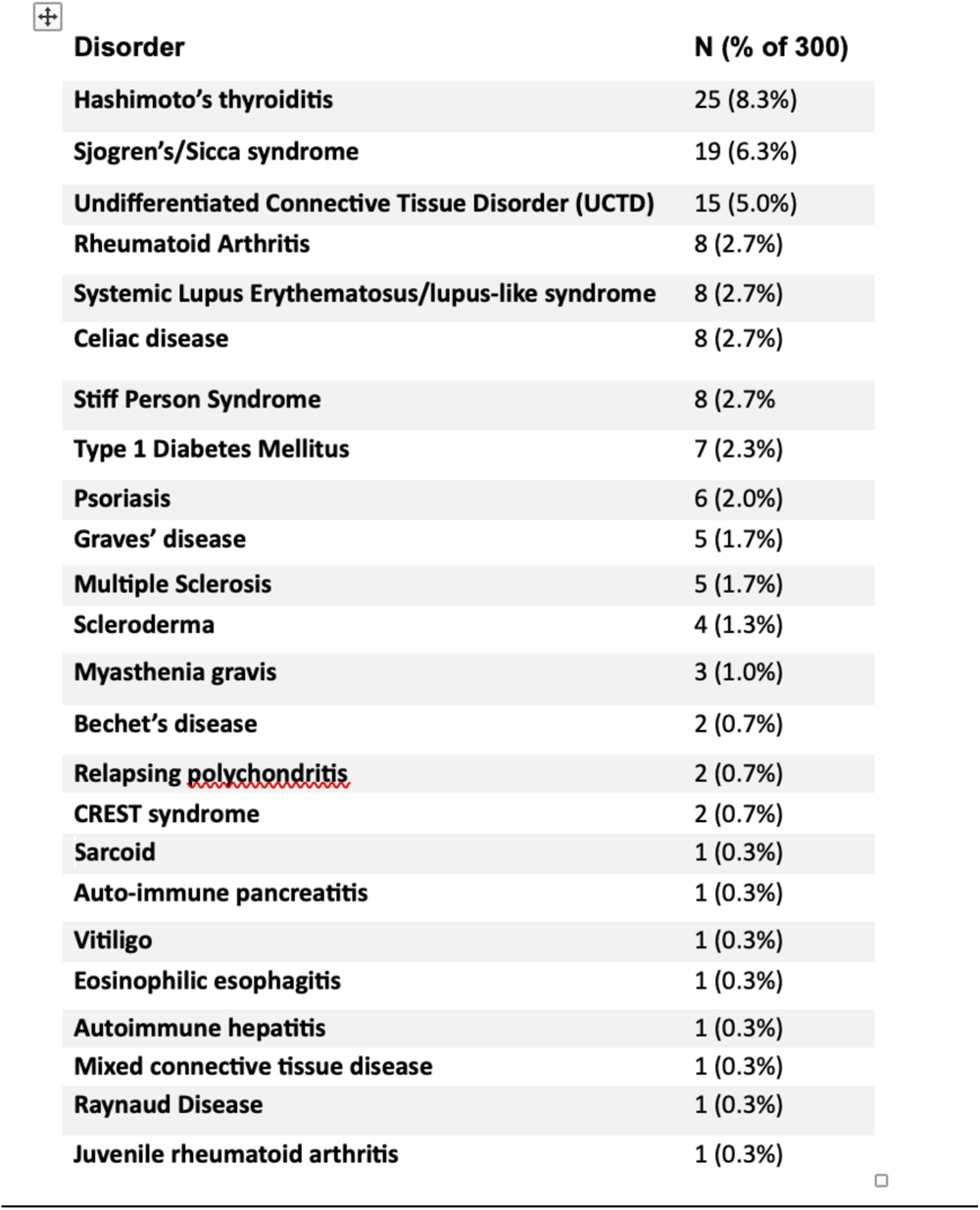
Autoimmune diagnoses in patients in retrospective cohort.

| Disorder | N (% of 300) |
| --- | --- |
| Hashimoto's thyroiditis | 25 (8.3%) |
| Sjogren's/Sicca syndrome | 19 (6.3%) |
| Undifferentiated Connective Tissue Disorder (UCTD) | 15 (5.0%) |
| Rheumatoid Arthritis | 8 (2.7%) |
| Systemic Lupus Erythematosus/lupus-like syndrome | 8 (2.7%) |
| Celiac disease | 8 (2.7%) |
| Stiff Person Syndrome | 8 (2.7%) |
| Type 1 Diabetes Mellitus | 7 (2.3%) |
| Psoriasis | 6 (2.0%) |
| Graves' disease | 5 (1.7%) |
| Multiple Sclerosis | 5 (1.7%) |
| Scleroderma | 4 (1.3%) |
| Myasthenia gravis | 3 (1.0%) |
| Bechet's disease | 2 (0.7%) |
| Relapsing polychondritis | 2 (0.7%) |
| CREST syndrome | 2 (0.7%) |
| Sarcoid | 1 (0.3%) |
| Auto-immune pancreatitis | 1 (0.3%) |
| Vitiligo | 1 (0.3%) |
| Eosinophilic esophagitis | 1 (0.3%) |
| Autoimmune hepatitis | 1 (0.3%) |
| Mixed connective tissue disease | 1 (0.3%) |
| Raynaud Disease | 1 (0.3%) |
| Juvenile rheumatoid arthritis | 1 (0.3%) |

As per the patients’ history, these disorders had been regarded as quiescent by the treating physician in most instances. An additional 22 patients in this cohort had a history of known autoimmune disorder but no associated antibody markers, and by our definition, not counted in the AIM group.

Autoantibody markers and their frequency is provided in Table 2; in patients who were ANA positive, the median titer was 1:160.

**Table 2:** Frequency of autoantibodies in retrospective cohort.

| **Autoantibody** | **Number of patients tested** | **Positive (n, %)** |
| --- | --- | --- |
| **Antiganglioside** | 219 | 79 (36.1) |
| **ANA** | 292 | 71 (24.3) |
| **ASMA** | 299 | 63 (21.1) * |
| **Anti-TPO** | 231 | 24 (10.3) |
| **Mayo Paraneoplastic Panel** | 279 | 23 (8.2) ** |
| **Anti-GAD65** | 273 | 19 (7.0) |
| **RF** | 285 | 6 (2.1) |
| **Anti-Scl70** | 279 | 3 (1.1) |
| **Anti-Smith** | 275 | 3 (1.1) |
| **Anti-Ro60** | 222 | 3 (1.3) |
| **Anti-DNA** | 280 | 2 (0.7) |
| **Anti-RNP** | 277 | 2 (0.7) |
| **Anti-Ro52** | 222 | 1 (0.5) |
| **Anti-SSB/La** | 213 | 0 (0.0) |

\*ASMA= Anti-Smooth Muscle Antibody; positive if titer >1:40
\*\*Striational (striated muscle) Ab = 5; Voltage-gated potassium channel Ab = 8; N-type calcium channel Ab = 1; AChR (acetylcholine receptor) ganglionic Ab = 6; AChR (muscle) binding Ab= 3 TPO = thyroid peroxidase

### Prospective cohort

#### Validation of retrospective results

Given the surprisingly high prevalence of AIM in the retrospective cohort, these results prompted us to initiate the prospective study to validate these results in a prospectively studied cohort as well to use standardized and published patient questionnaires to assess clinical correlates. A total of 133 patients were recruited in this cohort.

With respect to conventional gastrointestinal diagnoses based only on symptoms, these patients fulfilled criteria for multiple Rome IV categories (Table 3), totaling 566 in number.

**Table 3:** Rome IV classification of patients in prospective cohort (n=133)

| **ROME IV syndrome classification** | **N** |
| --- | --- |
| Functional Dyspepsia (FD) | 92 |
| FD/Postprandial Distress | 81 |
| FD/Epigastric Pain Syndrome | 60 |
| Belching Disorders | 17 |
| Nausea and Vomiting Disorders | 36 |
| Chronic Nausea and Vomiting Syndrome | 29 |
| Cyclic Vomiting Syndrome | 15 |
| Cannabinoid Hyperemesis | 1 |
| Rumination Syndrome | 20 |
| Bowel Disorders | 107 |
| Irritable Bowel Syndrome | 54 |
| Functional Constipation | 35 |
| Functional Diarrhea | 5 |
| Functional Abdominal Bloating/Distention | 3 |
| Unspecified Functional Bowel Disorder | 9 |
| Opioid-Induced Constipation | 2 |
| Centrally Mediated Abdominal Pain Syndrome | 0 |

Alternatively, some of these patients could also be given diagnoses based on the results of gut transit studies, performed in 102 patients of which 92 patients (90%) had delay in at least one organ. Fifty-four (53%) of these met criteria for gastroparesis (2-hour retention >60% and/or 4-hour retention >10%),^29^ 12 (12%) had delayed small bowel transit and 65 (64%) met criteria for slow transit constipation based on transit times previously published.^30^ In addition, 31 (30%) patients also had esophageal delay (even though dysphagia was not an obvious symptom in the vast majority).

Of the 133 patients, 47 (35%) met the pre-specified definition of autoimmunity, validating what was observed in the retrospective cohort (37%). Of the 47 patients, 27 (58%) patients had a history of a known rheumatic disorder, along with at least one autoantibody marker (Table 4). The top five rheumatic diagnoses in this group were the same as in the retrospective cohort namely, Hashimoto’s thyroiditis, Sjogren’s syndrome, Systemic Lupus Erythematosus (SLE)/lupus-like syndrome, Mixed/Undifferentiated connective tissue disorder and Rheumatoid Arthritis.

**Table 4.** Autoimmune diagnoses in patients in prospective cohort (n=133)

| **Disorder** | **N (% of 133 patients)** |
| --- | --- |
| Hashimoto's thyroiditis | 13 (9.8%) |
| Sjogren's/Sicca syndrome | 8 (6%) |
| Systemic Lupus Erythematosus/lupus-like syndrome | 6 (4.5%) |
| Mixed/Undifferentiated connective tissue disorder (MCTD/UCTD) | 3 (2.3%) |
| Rheumatoid Arthritis | 3 (2.3%) |
| Ankylosing Spondylitis | 3 (2.3%) |
| Celiac disease | 2 (1.5%) |
| Bechet's disease | 2 (1.5%) |
| Psoriasis | 1 (0.75%) |
| Graves' disease | 1 (0.75%) |
| Auto-immune pancreatitis | 1 (0.75%) |
| Pernicious anemia | 1 (0.75%) |

Autoantibody markers and their frequency is provided in Table 5: the five most prevalent antibodies in the prospective cohort were the same as that in the retrospective cohort-anti-ganglioside, anti-TPO, ANA, Anti-Smooth muscle antibody and the Mayo Paraneoplastic panel as a group.

**Table 5:** Autoantibodies in prospective cohort.

| **Autoantibody** | **Number of patients tested** | **Positive (n, %)** |
| --- | --- | --- |
| **Antiganglioside** | 86 | 23 (26.7) |
| **Anti-TPO** | 98 | 20 (20.4) |
| **ANA** | 114 | 20 (17.5) |
| **ASMA \*\*** | 39 | 5 (12.8) |
| **Mayo Paraneoplastic Panel \*** | 99 | 8 (8.1) |
| **Anti-SSA/Ro62** | 77 | 6 (7.8) |
| **Anti-SSB/La** | 49 | 3 (6.1) |
| **Anti-SSA/Ro52** | 77 | 4 (5.2) |
| **Anti-GAD65** | 101 | 4 (4.0) |
| **RF** | 105 | 4 (3.8) |
| **Anti-Scl70** | 100 | 1 (1.0) |
| **Anti-RNP** | 102 | 1 (0.98) |
| **Anti-Smith** | 103 | 1 (0.97) |

\*ASMA= Anti-Smooth Muscle Antibody; positive if titer >1:40
\*\*Striational (striated muscle) Ab = 2; Voltage-gated potassium channel Ab = 1; N-type calcium channel Ab = 1; AChR (acetylcholine receptor) ganglionic Ab = 3; Voltage-gated calcium channel = 1; Anti-GAD = 1
TPO = thyroid peroxidase

#### Phenotypic differences between patients with AIM and those without

Having validated that autoimmune markers are present in significantly large number of patients, we next took advantage of the prospective nature of this cohort and the use of validated patient reported outcomes to examine what, if any, were the differences in the clinical features and symptom severity as wellas demographical features (age, gender, ethnicity, and BMI) of patients with and without AIM. The results are shown in Table 6.

**Table 6.** Comparison of symptoms and demographics between patients with and ¥)without autoimmune markers (AIM)’

|  | **AIM**<br>**(N = 47)** | **No AIM**<br>**(N = 86)** | **$\Delta$** | **95% CI** | **P<sup>a</sup>** |
| --- | --- | --- | --- | --- | --- |
| **Female** | 42 (89%) | 66 (77%) | 0.13 | -0.02 to 0.27 | 0.122 |
| **Hispanic** | 2 (4%) | 3 (3%) | 0.01 | -0.07 to 0.08 | >0.999 |
| **Age (y)** | 44 (17) | 41 (17) | 2 (17) | -4 to 9 | 0.430 |
| **BMI** | 26 (6) | 24 (6) ** | 2 (6) | -1 to 4 | 0.155 |
| **JH/HSD** | 33 (70%) | 34 (40%) | 0.31 | 0.12 to 0.49 | 0.001<sup>b</sup> |
| **PAGI-SYM Total Score** | 2.4 (1.0) | 2.2 (1.0) | 0.2 (1.0) | -0.1 to 0.6 | 0.218 |
| **PAGI-QOL Total Score** | 2.7 (1.2) | 2.8 (1.1) | -0.1 (1.2) | -0.5 to 0.3 | 0.567 |
| **PAC-SYM Total Score** | 1.7 (0.9) | 1.5 (0.8) | 0.2 (0.8) | -0.1 to 0.5 | 0.107 |
| **PAC-QOL Total Score** | 2.2 (0.8) | 1.9 (0.9) ** | 0.3 (0.8) | 0.0 to 0.6 | 0.060 |
| **PFDI-20 Total Score** | 90 (70) | 60 (50) | 20 (60) | 0 to 50 | 0.019 |
| **COMPASS-31 Total Score** | 44 (20) | 36 (19) | 8 (20) | 1 to 15 | 0.019 |
| **Dysautonomia present<sup>c</sup>** | 34 (72%) | 50 (58%) | 0.14 | -0.04 to 0.32 | 0.151 |
| **MPQ PRI Total Score** | 16 (11) | 15 (11) | 1 (11) | -3 to 5 | 0.606 |
| **MPQ VAS** | 53 (25) | 45 (31) | 7 (29) | -3 to 18 | 0.161 |
| **MPQ PPI** | 1.7 (1.2) | 1.7 (1.5) | 0.0 (1.4) | -0.5 to 0.5 | 0.916 |
| **PROMIS Belly Pain Score** | 63 (13) | 60 (13) | 3 (13) | -2 to 7 | 0.273 |
| **PROMIS Pain Intensity** | 66 (11) | 62 (10) | 4 (10) | 0 to 8 | 0.035 |
| **PROMIS Pain Interference** | 62 (9) | 58 (11) | 4 (10) | 0 to 7 | 0.052 |
| **PROMIS Nociceptive Score** | 48 (10) | 45 (10) | 2 (10) | -1 to 6 | 0.203 |
| **PROMIS Neuropathic Score** | 44 (8) | 42 (8) | 2 (8) | -1 to 4 | 0.282 |
| **PROMIS Gas Score** | 62 (7) | 60 (9) ** | 2 (8) | -1 to 5 | 0.206 |
| **PROMIS Diarrhea** | 52 (10) | 51 (9) | 0 (10) | -3 to 4 | 0.806 |
| **PROMIS Incontinence** | 5.2 (2.6) | 5.2 (2.2) | 0.0 (2.3) | -0.8 to 0.8 | 0.988 |
| **PROMIS Physical Function** | 40 (9) | 43 (10) | -3 (9) | -6 to 0 | 0.073 |
| **PROMIS Anxiety** | 56 (10) | 57 (11) | -1 (11) | -5 to 2 | 0.468 |
| **PROMIS Depression** | 53 (11) | 55 (11) | -1 (11) | -5 to 3 | 0.536 |
| **PROMIS Fatigue** | 65 (12) | 61 (11) | 3 (11) | -1 to 7 | 0.105 |
| **PROMIS Sleep** | 58 (8) | 55 (9) | 2 (9) | -1 to 6 | 0.126 |
| **PROMIS Social** | 41 (11) | 44 (11) | -3 (11) | -7 to 1 | 0.166 |
| **HSQ-Migraine** | 5.1 (2.6) | 4.5 (2.3) | 0.6 (2.4) | -0.2 to 1.5 | 0.145 |
| **HSQ-TTH** | 5.9 (1.5) | 6.3 (1.4) | -0.4 (1.4) | -0.9 to 0.2 | 0.166 |

\*Mean (SD) or n (%)
\*\* n = 85
<sup>a</sup>Two-sample t test or Pearson chi-square test.
<sup>b</sup>5% FDR
<sup>c</sup>Defined by a COMPASS-31 score of >32
PAGI-SYM = patient assessment of upper gastrointestinal symptom severity index; PAGI-QOL = Patient Assessment of Upper Gastrointestinal Disorders-Quality of Life; PAC-SYM = Patient Assessment of Constipation-Symptoms; PAC-QOL = Patient Assessment of Constipation Quality of Life; PFDI = Pelvic Floor Distress Inventory; COMPASS = Composite Autonomic Symptom Score; MPQ = McGill Pain Questionnaire, PRI = Pain Rating Indices, VAS = Visual Analog Scale, PPI = Present Pain Intensity; HSQ = Headache Screening Questionnaire; JH/HSD = joint hypermobility/hypermobility spectrum disorder.

The most striking, (and the only statistically significant difference meeting 5% FDR criteria for multiple comparisons) amongst these two groups was in the prevalence of joint hypermobility/hypermobility spectrum disorder (JH/HSD) which was reported 70% of patients in the AIM-positive group versus only 40% in the AIM-negative group (nominal p =0.001). As defined by their COMPASS-31 score, the majority (84 out of 133 patients; 63%) of patients had dysautonomia but its prevalence was not different in the two groups. However, the severity of dysautonomia appeared to be higher in the AIM group, with a nominal P value of .019 but this difference did not meet the FDR threshold. The severity of pelvic floor symptoms (as measured on the PFDI scale) also was higher in the AIM group (nominal P of .019). Subscales of the measures used in this cohort and their contribution to the differences in the total scores can be found in the Appendix (Supplemental Table 1).

#### Prospective cohort: differences between patients with and without joint hypermobility

Given the remarkable representation of patients with JH/HSD in the AIM group, we further analyzed differences in the clinical presentation in patients with and without JH/HSD. After adjustment for multiple comparisons, several significant differences were found between these two groups (Table 7). 94% of patients with JH/HSD were female as compared with 68% in the group without this feature (P <.001). The prevalence of AIM was more than two-fold higher in the group with JH/HSD (49% versus 21%; P =.001). Upper gastrointestinal symptoms (PAGI-SYM) and associated quality of life (PAGI-QOL) were also worse in patients with JH/HSD, as were constipation related symptoms (PAC-SYM and PROMIS Gas) and quality of life (PAC-QOL). Dysfunctional pelvic floor symptoms (as measured by PFDI) were also remarkably higher in the JH/HSD group and may have contributed to the constipation severity. Pain scores, particularly as measured by PROMIS intensity and interference measures was also worse in patients with JH/HSD. HSQ-migraine but not -TTH scores, were also higher; in addition, 50 patients with JH/HSD (74.6%) had a score indicating “probable or definite migraine” (score of ≥ 6) as compared with 38 (57.6%) patients without JH/HSD (p = .045, Fisher’s exact test).^31^ Subscales of the measures used in this cohort and their contribution to the differences in the total scores can be found in the Appendix (Supplemental Table 2).

**Table 7.** Comparison of symptoms and demographics between patients with and without joint hypermobility/hypermobility spectrum disorder (JH/HSD) *.

| | JH/HSD<br>(N = 67) | Not JH/HSD<br>(N = 66) | $\Delta$ | 95% CI | $P^a$ |
| --- | --- | --- | --- | --- | --- |
| Female | 63 (94%) | 45 (68%) | 0.26 | 0.12 to 0.40 | <0.001<sup>b</sup> |
| Hispanic | 2 (3%) | 3 (5%) | -0.02 | -0.10 to 0.06 | 0.986 |
| Age (y) | 39 (17) | 45 (16) | -6 (17) | -12 to 0 | 0.035 |
| BMI | 24 (6) | 25 (7)** | -2 (6) | -4 to 1 | 0.142 |
| Autoimmune markers | 33 (49%) | 14 (21%) | 0.28 | 0.11 to 0.45 | 0.001<sup>b</sup> |
| PAGI-SYM Total Score | 2.5 (0.9) | 2.1 (1.0) | 0.4 (0.9) | 0.1 to 0.8 | 0.007<sup>b</sup> |
| PAGI-QOL Total Score | 2.6 (1.2) | 3.0 (1.1) | -0.4 (1.1) | -0.8 to 0.0 | 0.063 |
| PAC-SYM Total Score | 1.8 (0.9) | 1.3 (0.7) | 0.5 (0.8) | 0.2 to 0.7 | 0.001<sup>b</sup> |
| PAC-QOL Total Score | 2.2 (0.9) *** | 1.8 (0.8) | 0.4 (0.8) | 0.1 to 0.7 | 0.005<sup>b</sup> |
| PFDI-20 Total Score | 85 (62) | 57 (52) | 28 (57) | 9 to 48 | 0.005<sup>b</sup> |
| COMPASS-31 Total Score | 47 (19) | 31 (18) | 16 (18) | 10 to 22 | <0.001<sup>b</sup> |
| Dysautonomia present | 55 (82%) | 29 (44%) | 0.38 | 0.22 to 0.55 | <0.001<sup>b</sup> |
| MPQ PRI Total Score | 18 (11) | 13 (11) | 4 (11) | 0 to 8 | 0.033 |
| MPQ VAS | 52 (27) | 44 (31) | 8 (29) | -2 to 18 | 0.112 |
| MPQ PPI | 1.8 (1.3) | 1.6 (1.5) | 0.2 (1.4) | -0.3 to 0.7 | 0.400 |
| PROMIS Belly Pain Score | 64 (12) | 59 (14) | 5 (13) | 1 to 9 | 0.028 |
| PROMIS Pain Intensity | 66 (10) | 61 (11) | 5 (10) | 2 to 9 | 0.004<sup>b</sup> |
| PROMIS Pain Interference | 61 (10) | 57 (10) | 4 (10) | 0 to 7 | 0.025<sup>b</sup> |
| PROMIS Nociceptive Score | 47 (10) | 45 (10) | 2 (10) | -1 to 6 | 0.233 |
| PROMIS Neuropathic Score | 43 (8) | 42 (8) | 1 (8) | -1 to 4 | 0.322 |
| PROMIS Gas Score | 63 (7) *** | 59 (8) | 3 (8) | 0 to 6 | 0.021<sup>b</sup> |
| PROMIS Diarrhea | 52 (9) | 51 (10) | 1 (10) | -2 to 4 | 0.472 |
| PROMIS Incontinence | 5.2 (2.5) | 5.2 (2.1) | 0.1 (2.3) | -0.7 to 0.9 | 0.830 |
| PROMIS Physical Function | 40 (10) | 44 (9) | -4 (9) | -7 to -1 | 0.021<sup>b</sup> |
| PROMIS Anxiety | 58 (10) | 56 (11) | 2 (11) | -1 to 6 | 0.216 |
| PROMIS Depression | 55 (11) | 53 (11) | 3 (11) | -1 to 6 | 0.179 |
| PROMIS Fatigue | 65 (11) | 60 (11) | 4 (11) | 1 to 8 | 0.021<sup>b</sup> |
| PROMIS Sleep | 58 (8) | 54 (9) | 4 (9) | 1 to 7 | 0.010<sup>b</sup> |
| PROMIS Social | 41 (11) | 45 (10) | -5 (11) | -8 to -1 | 0.014<sup>b</sup> |
| HSQ-Migraine | 5.4 (2.2) | 4.1 (2.4) | 1.3 (2.3) | 0.5 to 2.1 | 0.002<sup>b</sup> |
| HSQ-TTH | 6.1 (1.3) | 6.2 (1.6) | 0.0 (1.5) | -0.5 to 0.5 | 0.851 |

\*Mean (SD) or n (%);
\*\* n = 65; \*\*\* n = 66
<sup>a</sup> Two-sample t test or Pearson chi-square test.
<sup>b</sup> 5% FDR
<sup>c</sup> Defined by a COMPASS-31 score of >32
PAGI-SYM = patient assessment of upper gastrointestinal symptom severity index; PAGI-QOL = Patient Assessment of Upper Gastrointestinal Disorders-Quality of Life; PAC-SYM = Patient Assessment of Constipation-Symptoms; PAC-QOL = Patient Assessment of Constipation Quality of Life; PFDI = Pelvic Floor Distress Inventory; COMPASS = Composite Autonomic Symptom Score; MPQ = McGill Pain Questionnaire, PRI = Pain Rating Indices, VAS = Visual Analog Scale, PPI = Present Pain Intensity; HSQ = Headache Screening Questionnaire

Even more striking than the differences in gastrointestinal symptoms, was the severity of dysautonomia in JH/HSD group, with a mean COMPASS-31 score that was 16 points higher than the group without JH/HSD. To put this in perspective, we compared the scores in these two patient groups with data reported elsewhere on healthy volunteers, patient with diabetic polyneuropathy, small fiber neuropathy and scleroderma.^32^ The results are displayed in Figure 1; as can be seen, patients with JH/HSD had a significantly higher score than patients without JH/HSD as well as those with other well-known neurological disorders.

**Figure 1.**
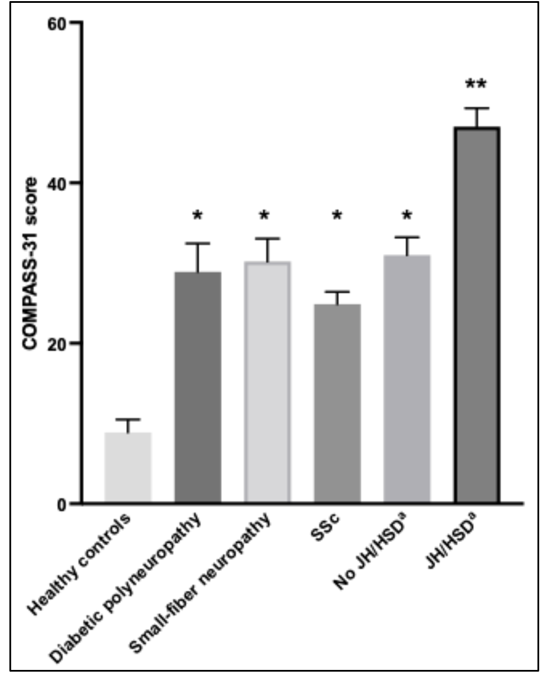
COMPASS-31 scores (Means + S.E.M.), a measure of autonomic dysfunction, of patients with JH/HSD and without^a^, as compared with healthy volunteers and patients with other disorders. *Compass scores in all disease categories were significantly higher than healthy controls (P < .001, oneway ANOVA). **Compass scores in patients with JH/HSD were significantly higher than all other disease categories (P<0.001). Compass scores in all other disease categories, including patients without JH/HSD were not significantly different than each other. SSC= Systemic scleroderma; JH/HSD = joint hypermobility/Hypermobility Spectrum Disorder a Patients in this study. Other data from a previously published report and provided for comparison.25

#### Association between autoimmunity and objective markers of disease

Next, we attempted to examine whether laboratory tests could help distinguish patients with AIM from those without. Since these were objective results, we pooled the data from both prospective and retrospective cohorts to increase statistical power, giving a total of 433 patients (Table 8). In addition to blood tests that were done in almost all patients, skin biopsies were done in 180 patients (of whom 31% had small fiber neuropathy), provocative cardiovascular tilt table testing was done in 210 patients (of whom 74% had a positive test for Postural Orthostatic Tachycardia Syndrome/POT and/or Neurally Mediated Hypotension/NMH) and whole gut scintigraphy in 366 patients (of whom 74% had delayed colonic transit).

**Table 8.** Comparison of objective tests and demographics between patients with and without autoimmune markers (AIM)*

| | AIM<br>(N = 164) | No AIM<br>(N = 269) | $\Delta$ | 95% CI | *P*<sup>a</sup> |
| --- | --- | --- | --- | --- | --- |
| Female | 148 (90%) | 226 (84%) | 0.06 | -0.01 to 0.13 | 0.091 |
| Age (y) | 43 (16) | 40 (17) | 3 (17) | 0 to 6 | 0.079 |
| CRP High | 51/158 (32%) | 36/252 (14%) | 0.18 | 0.09 to 0.27 | <0.001<sup>b</sup> |
| Sed Rate High | 41/158 (26%) | 23/248 (9%) | 0.17 | 0.08 to 0.25 | <0.001<sup>b</sup> |
| IgA Low | 16/156 (10%) | 41/230 (18%) | -0.08 | -0.15 to 0.00 | 0.056 |
| IgA High | 5/156 (3.2%) | 9/230 (3.9%) | -0.007 | -0.050 to 0.036 | 0.930 |
| IgG Low | 16/162 (10%) | 28/246 (11%) | -0.02 | -0.08 to 0.05 | 0.752 |
| IgG High | 7/162 (4.3%) | 5/246 (2.0%) | 0.023 | -0.018 to 0.064 | 0.299 |
| IgM Low | 6/156 (4%) | 13/230 (6%) | -0.02 | -0.07 to 0.03 | 0.572 |
| IgM High | 13/156 (8%) | 11/230 (5%) | 0.04 | -0.02 to 0.09 | 0.229 |
| IgG1 Low | 35/158 (22%) | 41/237 (17%) | 0.05 | -0.04 to 0.13 | 0.285 |
| IgG1 High | 6/158 (3.8%) | 2/237 (0.8%) | 0.030 | -0.008 to 0.067 | 0.094 |
| IgG2 Low | 37/158 (23%) | 53/237 (22%) | 0.01 | -0.08 to 0.10 | 0.903 |
| IgG2 High | 5/158 (3.2%) | 3/237 (1.3%) | 0.019 | -0.017 to 0.055 | 0.343 |
| IgG3 Low | 16/158 (10%) | 24/237 (10%) | 0.00 | -0.06 to 0.06 | >0.999 |
| IgG3 High | 2/158 (1.3%) | 0/237 (0.0%) | 0.013 | -0.010 to 0.035 | 0.311 |
| IgG4 Low | 9/158 (6%) | 12/238 (5%) | 0.01 | -0.04 to 0.06 | 0.956 |
| IgG4 High | 8/158 (5%) | 20/238 (8%) | -0.03 | -0.09 to 0.02 | 0.285 |
| Small Fiber Neuropathy | 32/86 (37%) | 24/94 (26%) | 0.12 | -0.03 to 0.26 | 0.126 |
| Sudomotor Neuropathy | 11/80 (14%) | 7/87 (8%) | 0.06 | -0.05 to 0.16 | 0.348 |
| Tilt Table | 76/93 (82%) | 79/117 (68%) | 0.14 | 0.02 to 0.27 | 0.030 |
| Tryptase | 7/132 (5%) | 6/190 (3%) | 0.02 | -0.03 to 0.07 | 0.500 |
| tTG-IgA | 4/145 (2.8%) | 2/213 (0.9%) | 0.018 | -0.017 to 0.054 | 0.370 |
| Celiac HLA DQ2 | 33/148 (22%) | 53/223 (24%) | -0.01 | -0.11 to 0.08 | 0.839 |
| Celiac HLA DQ8 | 35/149 (23%) | 29/223 (13%) | 0.10 | 0.02 to 0.19 | 0.013 |
| Gastric Delay | 54/147 (37%) | 76/230 (33%) | 0.04 | -0.07 to 0.14 | 0.532 |
| Small Bowel Delay | 15/145 (10%) | 22/223 (10%) | 0.00 | -0.06 to 0.07 | >0.999 |
| Colon Delay | 105/145 (72%) | 165/221 (75%) | -0.02 | -0.12 to 0.08 | 0.721 |
| Delays >= 2 (of 3) | 50/144 (35%) | 62/220 (28%) | 0.07 | -0.04 to 0.17 | 0.228 |

\* Mean (SD) or n (%).
<sup>a</sup> Two-sample *t* test or Pearson chi-square test.
<sup>b</sup> 5% FDR
tTG-IgA = tissue transglutaminase (IgA)

The largest differences between groups with and without AIM were for high C-reactive protein (CRP) and high erythrocyte sedimentation rate or ESR (Table 8). High CRP was present in 32% of patients with autoimmunity versus only 14% in those without autoimmunity, and high ESR was present in 26% of patients with autoimmunity versus only 9% of those without autoimmunity, both of these differences were significant (P values less than 0.002 indicated 5% FDR with correction for 29 comparisons). Although no other variables met this criterion, a positive tilt-table test (83% versus 68%) and HLA DQ8 (23% versus 13%) were higher in patients with AIM, with nominal p-values of .03 and .01, respectively.

#### IVIG treatment results in significant clinical improvement in multiple gastrointestinal and autonomic symptoms

Forty-two (42) patients suspected to have autoimmune disease with gastrointestinal complaints refractory to both standard and off-label symptomatic therapies (see Table 9 for a representative list of medications that these patients received) were treated with IVIG by a single physician and followed prospectively.

**Table 9:**
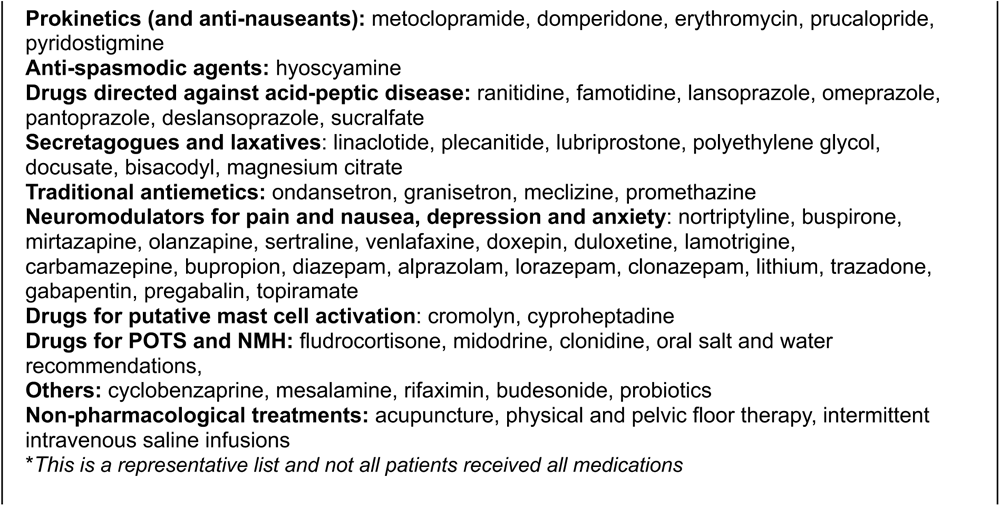
List of Medications Patients Were Tried on Before Considering IVIG*.

Eight patients were excluded because they either did not complete more than 2 courses (because of lack of insurance coverage in 4 and adverse effects in 2) or did not complete the surveys (n=2). Of the remaining 34 patients, 32 had AIM, had undergone at least 3 courses of IVIG with completed surveys and were included for analysis.

Twenty-nine (91%) of the 32 patients treated with IVIG, ranged in age from 18 to 76 years, with a mean of 35 years. JH/HSD was present in 75% of patients. The commonest antibody markers were against gangliosides (13/24; 54%), ANA in 12/32;38%), smooth muscle antigen (8/31; 26%), thyroid peroxidase (5/22; 23%) and the Mayo paraneoplastic panel (6/31;19%). Gut transit studies showed gastric delay (gastroparesis) in 33% (10/30), small intestinal delay in 3.4% (1/29), and colonic delay in 83% (24/29), with 32% having delays of more than one of these regions. Tilt table was positive in 81% (22/27); skin biopsy (n = 31) showed small fiber and sudomotor neuropathy in 16% (5/31) and 6% (2/31), respectively.

Overall treatment effectiveness (OTE) scores as assessed on a 15-point scale from -7 to +7, are shown in Figure 2. The mean was significantly better than zero (95% CI 0.6 to 2.9, *P* 0.004). The mean was more than 25% of the magnitude of the scale, and the effect size was more than 0.5 standard deviations (indicating at least a medium effect). Scores ranged from -5 to 7 with a mean of 1.8 (SD 3.2); 16 patients (50%) reported a score of at least 3 (somewhat better) and 10 (31%) reported a score of at least 4 (moderately better). Validated patient reported outcomes such PAGI-SYM, PAGI-QOL, GCSI, PAC-QOL, PFDI-20, Eckardt, and COMPASS-31 scores were markedly improved in the patients that received IVIG (Table 10).

**Figure 2.**
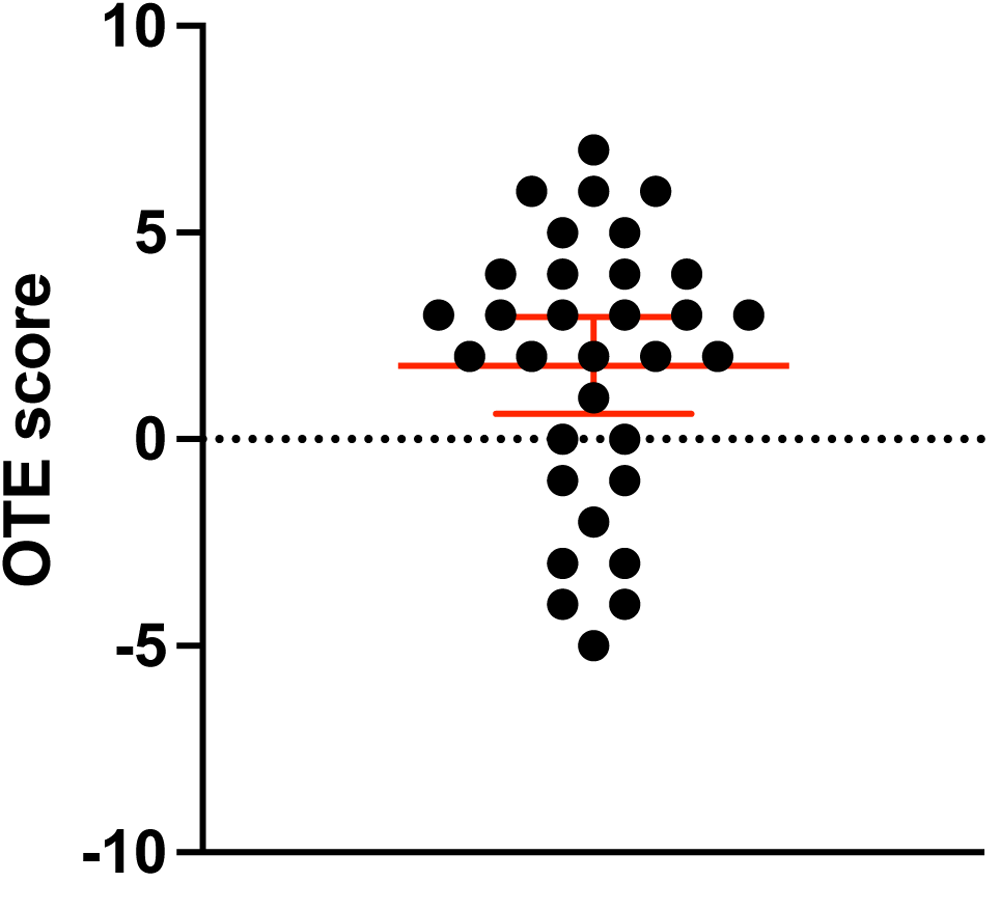
Distribution of overall treatment effect (OTE) scores following treatment with IVIG. Scatter dot plot of individual responses with mean and 95% CI indicated in red.

**Table 10.** Change in patient reported outcome scales and subscales following IVIG treatment.

| Variable | N | Pre | Post | ----- Change ----- |  |  |
| --- | --- | --- | --- | --- | --- | --- |
|  |  | Mean (SD) | Mean (SD) | Mean (SD) | 95% CI | *P*<sup>a</sup> |
| **PAGI-SYM** | 32 | 2.7 (1.0) | 2.0 (1.1) | -0.7 (0.9) | -1.1 to -0.4 | <0.001<sup>b</sup> |
| **PAGI-QOL** | 32 | 1.9 (1.0) | 2.8 (1.2) | 0.9 (0.9) | 0.5 to 1.2 | <0.001<sup>b</sup> |
| **PAC-SYM** | 9 | 1.8 (1.3) | 1.1 (0.9) | -0.8 (1.1) | -1.6 to 0.1 | 0.073 |
| **PAC-QOL** | 32 | 2.2 (1.1) | 1.7 (0.8) | -0.5 (0.8) | -0.8 to -0.2 | 0.001<sup>b</sup> |
| **PFDI-20** | 32 | 95 (49) | 76 (45) | -19 (38) | -32 to -5 | 0.008<sup>b</sup> |
| **Eckardt** | 31 | 3.5 (3.0) | 2.3 (2.3) | -1.2 (2.3) | -2.0 to -0.3 | 0.007<sup>b</sup> |
| **COMPASS-31** | 12 | 58 (17) | 41 (14) | -17 (12) | -25 to -10 | <0.001<sup>b</sup> |

<sup>a</sup> Paired *t* test.
<sup>b</sup> Hochberg *P* < 0.050.

The largest effect size was observed for the COMPASS-31 (ES 1.5), but scores were only available for 12 patients. The PAGI-QOL (Figure 2) had the largest effect size among scales measured for all subjects (ES 0.9). Quality of life, as measured by PAGI-QOL, improved mean scores by 0.9, which corresponds to a “great or very great deal better” ^16^; mean scores on PAC-QOL also improved by 0.5, meeting the threshold for a clinically meaningful response.^18^ Subscales of the measures used in this cohort and their contribution to the differences in the total scores can be found in the Appendix (Supplemental Table 3).

The sample size was too small to allow for any meaningful analysis of predictors of response. However, it should be noted that 80% of patients with an Overall Treatment Effect score of “3” or higher had Other Autoimmune Diagnosis versus 43% among those with a score less than “3” (OR 5.3, P 0.094).

Adverse events from IVIG were very common, occurring in 65% of all treated patients. Headaches were the most frequent and serious side-effect, reported by 13 patients (39%); of these, 6 patients presented to the emergency room with severe headaches, 4 (12% of all patients) of whom were diagnosed with aseptic meningitis. Other adverse events included skin rash (n = 4; 12%), nausea/vomiting (n = 3; 9%), edema/weight gain/bloating (n =3; 9%), fever/chills (n = 2; 6%), transient hypertension (n =2; 6%), chest tightness/shortness of breath (n = 2; 6%), back pain, fatigue, abnormal taste in mouth, diffuse muscle spasms, abdominal pain, loss of appetite (n= 1 each; 3%).

## Discussion

The identification of novel autoimmune diseases usually occurs through extensive research, clinical observations, and diagnostic investigations. This study can be considered as part of the initial steps in this process by studying the association of autoimmune markers in a large population of carefully phenotyped patients with severe but unexplained gastrointestinal symptoms. These patients came to us with complaints of a combination of abdominal pain and disturbances in gastrointestinal motility but were otherwise unselected. Although this was their primary presentation, we also sought and found evidence for disturbances in the autonomic nervous system.

Our findings are remarkable in several ways. First, we found a surprisingly high (35-40%) proportion of patients with autoimmune markers (AIM) that was confirmed in both the initial and the validation cohort. A diagnosis of an autoimmune disorder is not controversial in the more than half of these patients (73% in the retrospective cohort and 58% in the validation cohort) who had a history of a well-accepted autoimmune diagnosis, in addition to having at least one autoantibody. However, in almost all instances this diagnosis was thought to be quiescent or controlled by the referring physician(s) and not contributing to the present illness. To some extent, this position has been reinforced by the literature that reports a high prevalence and severity of gastrointestinal symptoms in conditions such as Sjogren’s but typically attributes them to irritable bowel syndrome or a related condition.^33^ In the minority of patients (in whom there was no previous diagnosis of a classic rheumatic disorder), we used the presence of at least two auto-antibodies to classify them as possible autoimmune in nature, which we recognize may be more controversial. Hence, we have used the term “autoimmune markers” (AIM) in this study for both groups rather than making a definitive statement that what we have described here reflects a classically proven autoimmune disorder.

Nevertheless, the group with AIM had a 2 to 3-fold greater prevalence of elevated CRP and ESR values, a finding that was highly significant and supportive of an underlying inflammatory disorder. Further, these patients were more likely to express HLA-DQ8 and although the nominal P value of .01 was not significant after correction for multiple comparisons, it cannot be completely ignored in the context of putative autoimmunity. HLA-DQ8 has been associated with several other autoimmune disorders (apart from celiac disease), including rheumatoid arthritis^34^, autoimmune thyroiditis^35^, T1 diabetes mellitus^36^, and Addison’s disease.^37^ Many of these disorders were present in our patients (Tables 1 and 3), which could account for this finding. Finally, tilt-table testing revealed evidence of POTS or NMH in a higher proportion of patients with AI (83% versus 68%, p = .03), suggesting a systemic targeting of peripheral nerves such as can be seen with autoimmunity. We believe that these results provide at least partial validation of our criteria for AIM positivity, reinforcing the notion that this may have some pathological significance.

Not unexpectedly, we found common autoantibodies such as antinuclear antibody (ANA), anti-thyroid peroxidase (anti-TPO) in addition to a variety of rarer antibodies. However, it should be noted that two of the most prevalent antibodies-anti-ganglioside antibodies and anti-smooth muscle antibodies (ASMA) have generally not been reported in this context. Anti-ganglioside antibodies have classically been described in Guillain-Barre Syndrome (GBS) and its variants that affect the peripheral nervous system, and may represent an immune reaction to *Campylobacter jejuni* whose lipopolysaccharides resemble gangliosides (molecular mimicry).^38^ ASMA, directed against actin, have generally been associated with autoimmune hepatitis, although they have been reported in up to 60% of patients with Sjogren’s, another condition that is commonly associated with autonomic and gastrointestinal symptoms^39, 40^ and often without overt liver disease.^41^ Our findings therefore suggest that both antiganglioside antibodies and ASMA may be associated with autoimmune peripheral neuropathy affecting the autonomic and enteric nervous systems, but we acknowledge that causality remains to be established. It should also be noted that the prevalence of AIM in our patient groups may be significantly underestimated as we used a relatively large, but still limited, set of commercially available autoantibodies; further, we cannot rule out the possibility of as yet unknown/unreported autoantibodies.

It can rightfully be argued that the patients described in our study represent a severe end of the spectrum due to the tertiary nature of the practice. However, other investigators have also reported evidence of autoimmunity in more “typical” patients with GI motility disorders. A larger prospective study of 78 patients meeting Rome III criteria for IBS reported that 87% of patients sera (compared with 59% of control sera) contained “anti-enteric neuronal” antibodies directed against nuclear antigens expressed by the guinea pig ENS, which were subsequently identified as various ribonucleoproteins including Ro-52, implicated in Sjogren’s syndrome and others.^42^. In another study of 10 patients meeting Rome I criteria for IBS, laparoscopically obtained full thickness jejunal biopsies showed intra-or peri-ganglionic infiltration of lymphocytes in the myenteric plexus of 9, six of which also displayed neuronal degeneration.^43^ These changes may be more widespread in patients with “functional” GI disorders than generally recognized because of limitations in obtaining tissue in those with mild to moderate symptoms and ready dismissal of their illness as a “functional/gut-brain disorder”.

Our study adds new information to the literature on a possible autoimmune etiology affecting the peripheral nervous systems. The most well described syndrome of autoimmunity that affects both the autonomic and enteric nervous systems is autoimmune autonomic ganglionopathy (AAG), defined by the detection of antibodies to the ganglionic acetylcholine receptor (gAChR, at levels greater than 0.20 nmol/L) in the context of diffuse failure of the sympathetic, parasympathetic and enteric nervous systems.^44^ This is a relatively rare disorder whose specificity appears to reside in the pathogenic nature of the antibody, shown by animal transfer studies and correlation of antibody titers with overall severity of disease. Lower levels of gAChR antibody are not uncommon in patients with both suspected gastrointestinal dysmotility (9%)^45^ as well as specific syndromes such as achalasia (21%) and chronic intestinal pseudo-obstruction (50%),^46^ and have been considered “non-specific”.^44^ However, this view has been challenged by recent descriptions of what has been called autoimmune gastrointestinal disease (AGID).^1, 47^ In a small study, 24 patients with dysmotility tested positive on a panel of antibodies against neuronal antigens (generally called the “Mayo paraneoplastic panel” collectively).^47^ Although these patients bear some resemblance to our study subjects, as a group they were older with less female predominance and most importantly, 11 patients had an underlying neoplasia which our patients did not.

Nearly three-quarters of our patients had objective delay in colon transit, and about a third had gastric emptying delay/gastroparesis but notably, more than 60% of all patients in the prospective group had dysautonomia. Multiple studies in the literature have described the prevalence of a variety of autoantibodies in POTS and related conditions, and an autoimmune etiology is likely in at least a subset of patients with these disorders.^48–51^ The prevalence or severity of gastrointestinal symptoms is not consistently reported in these studies but ranges from around 20-50%.^48, 50^ In this regard, our results may be seen as similar to what has been described in patients presenting with POTS/NMH. However, our study is distinguished by the fact that the predominant symptom affecting these patients was gastrointestinal in nature and most patients were not previously suspected to have autonomic dysfunction. There have been few systematic reports examining peripheral nerves in patients with GI dysmotility. In a small study of 33 patients with slow transit constipation, about a third were found to have reduced axon-reflex sweating and a similar proportion had small sensory fiber dysfunction using thermal threshold testing.^52^ In our study, about 74% of 210 patients who underwent tilt table testing (36% of all 433 patients) were diagnosed with postural orthostatic tachycardia syndrome (POTS) and/or neurally mediated hypotension (NMH) and amongst those who underwent skin biopsy, 31% (13% of all patients) had evidence of small fiber neuropathy and 11% (4% of all patients) had evidence of sudomotor/sweat gland neuropathy. Our results therefore underline the importance of maintaining a high state of vigilance for a systemic neuropathic condition that may involve the enteric, autonomic, and peripheral nervous systems in patients presenting with chronic unexplained gastrointestinal symptoms,

Importantly, this study also examined the relationship between joint hypermobility (JH) and gastrointestinal and/or autonomic symptoms. Beginning with the seminal study by Hakim and Grahame,^53^ there is increasing recognition of the involvement of the gastrointestinal (GI) tract and autonomic nervous system with joint hypermobility syndromes, supported by prospective broad-based surveys of this population of patients.^54, 55^ To diagnose JH, we used a self-reported instrument in the form of a five-part questionnaire (5PQ) that includes five aspects of past or present hypermobility, with a cut-off level of two positive answers to any of the five questions.^22^ It has been validated for use in patient cohorts in the clinic^56^ as well as population-based studies.^57^ It advantages over the Beighton score is that it does not require a physical examination, checks for previous agility, and therefore provides a more generalized assessment of hypermobility, rather than focusing exclusively on 5 joints. False negatives are expected to be less with this approach and its use has been accepted as an alternative way to identify patients with joint hypermobility.^23^ Because we did not evaluate these patients with the more stringent criteria required to make a diagnosis of hypermobile Ehler Danlos Syndrome (hEDS) we therefore use the preferred term “hypermobility spectrum disorders” (HSD) for these cases.^23^

It appears that JH/HSD, used either as an isolated feature (as in our study) or in combination with more rigid criteria, is a risk factor for gastrointestinal dysfunction (as well as multiple other co-morbidities including dysautonomia), occurring in a third to as many of three-quarters of patients, as comprehensively reviewed elsewhere.^58, 59^ Half of our patients had JH/HSD and these were significantly younger and more likely to be female, which is similar to what has been reported by others.^60,61^ However, a major contribution of this study over others in the literature has been to quantify the severity of gastrointestinal and autonomic symptoms using validated scores. Our results show that symptoms arising from the peripheral nervous systems are significantly more intense in patients with JH/HSD. The severity of dysautonomia (as measured by the COMPASS score) was markedly higher than that reported in other disorders of the peripheral nervous system (Figure 1). Pain (including migraine, but not tension-type headache) was also prominent using a variety of indices, but interestingly it is the sensory component (according to the McGill Pain Questionnaire, Supplemental Table 2) that is more intense in patients with JH/HSD (there is no difference in the affective component), indicating a disorder of pain signaling rather than emotional handling. Further, none of our patients met the Rome IV diagnostic criteria for centrally mediated abdominal pain syndrome (CAPS), widely believed to be the cause of pain in the kinds of patients that we describe here.^62^ Tellingly, the ability to participate in social roles and activities (as shown by the PROMIS social scores, Table 7) was more impaired in patients with JH/HSD, attesting to the impact of symptom severity on their day-to-day life. At the same time, measures of anxiety and depression did not differ and were modest in both groups and should lend support to the belief that these symptoms are not a result of psychological or primarily “brain-gut axis interaction” co-morbidities.

One of the most remarkable findings in this study was that almost 50% of patients with JH/HSD had AIM, an approximately 2.5-fold increase over those without JH/HSD (Table 7); conversely 70% of patients with AIM had JH/HSD as compared to 40% without (Table 6). Such an association has not been described before and adds to the myriad co-morbidities noted in patients with JH/HSD. Our results therefore provide further evidence that patients with JH/HSD appear to be predisposed to a disorder that affects three peripheral nervous systems (somatosensory, autonomic, and enteric)-a neural “triopathy”. Despite progress in our understanding of the clinical spectrum of JH/HSD, little is known about the pathogenesis of autonomic or gastrointestinal symptoms, although a genetic abnormality in the composition of the extracellular matrix is undoubtedly important, as shown by preclinical studies in mouse models.^63, 64^ However, the prevalence of JH/HSD in the general population is probably much higher than those that become symptomatic,^65^ suggesting the possibility of a ‘second-hit” that is acquired after birth. The strong association with AIM suggests that autoimmunity may be one such factor, perhaps triggered by an infection or other environmental factors. In this context it can be speculated that patients with joint hypermobility are predisposed to autoimmunity because of aberrant transforming growth factor ý (TGFý) signaling that may contribute to both connective tissue abnormalities and immune dysfunction, as has been reported in similar disorders in the literature.^66^ The importance of our finding, if validated by other studies, lies in the potential ability to intervene in a subset of patients with immunomodulatory regimens as described in this report.

To test the association of AIM with a response to immunomodulators, we conducted a prospective proof-of-concept trial of intravenous immunoglobulin in 32 patients that were followed by validated symptom questionnaires. IVIG resulted in significant improvements in OTE scores, as well as in validated quality of life measures for both upper (PAGI-QOL) and lower (PAC-QOL) and highly significant improvements in multiple gastrointestinal, autonomic symptoms and importantly, abdominal pain. However, it should be noted that IVIG treatment can have significant adverse events, with the incidence of headache and aseptic meningitis apparently higher in this population than that reported in general.^48^ Although the number of treated patients was too small to yield any statistically significant predictor, patients with a known autoimmune disorder did show a trend towards a better outcome than those who did not have such a history. Several of these disorders (e.g., hypothyroidism that is often due to Hashimoto’s thyroiditis, Sjogren’s, UCTD or psoriasis) are commonly present in patients but seldom if at all, considered as a cause of the gastrointestinal symptoms via a putative autoimmune enteric neuropathy, although they have been implicated in patients with POTS.^48^ Our results, which need to be validated by controlled trials (perhaps using other immunomodulators as well), will hopefully make physicians reconsider the importance of such diagnoses in patients presenting with otherwise unexplained gastrointestinal and/or autonomic disorders.

There are several limitations of this study that we acknowledge, including the tertiary nature of the practice, and the lack of a randomized controlled trial design for IVIG treatment. The strengths of this study include a prospective cohort that was administered multiple validated questionnaires, the large numbers of patients that were systematically studied using a comprehensive set of objective tests to measure whole gut transit and autonomic neuropathy (including tilt table and skin biopsies), a single gastroenterologist with a uniform clinical approach and the largest prospectively conducted trial of IVIG for gastrointestinal symptoms.

In conclusion, we describe four features that are commonly found in patients with unexplained gastrointestinal symptoms: joint hypermobility, <u>a</u>utonomic dysfunction, <u>g</u>astrointestinal dysfunction and <u>a</u>utoimmune markers; we have termed these features collectively as “JAGA”, if all four are present; however, patients can also present with various combinations of these features. Patients with joint hypermobility appear to be particularly prone to display autoimmune markers and dysautonomia (in addition to having more severe gastrointestinal symptoms and pain). It is therefore important for gastroenterologists to screen for joint hypermobility, autonomic dysfunction and autoimmune markers in patients presenting with unexplained gastrointestinal symptoms. Gastrointestinal and autonomic symptoms in these patients, particularly those with a history of known autoimmune disease such as thyroiditis, may respond to immunomodulation, as suggested by our initial findings. However, these hypotheses need to need to be validated by more robustly controlled studies, coupled with further research to establish a true autoimmune disorder in these patients.

## Data Availability

All data produced in the present work are contained in the manuscript

## Acknowledgement

We gratefully acknowledge the help of Dr. Gayane Yenokyan, MD, MPH, PhD, Executive Director, Johns Hopkins Biostatistics Center, for statistical help during the initial analysis.

There are no conflicts of interest for any of the authors.

## APPENDIX: SUPPLEMENTARY TABLES

**Supplemental Table 1.** Comparison of symptom subscales between patients with and without autoimmune markers (AIM)*

Terms in bold indicate the global test scores (displayed in Table 6)
|  | **AI**<br>**(N = 47)** | **Not AI**<br>**(N = 86)** | **$\Delta$** | **95% CI** | **P<sup>a</sup>** |
| --- | --- | --- | --- | --- | --- |
| **PAGI-SYM** |  |  |  |  |  |
| Nausea Score | 1.4 (1.1) | 1.5 (1.3) | -0.1 (1.3) | -0.6 to 0.3 | 0.533 |
| Fullness Score | 2.9 (1.3) | 2.8 (1.4) | 0.2 (1.3) | -0.3 to 0.7 | 0.436 |
| Bloating Score | 3.3 (1.4) | 2.8 (1.7) | 0.4 (1.6) | -0.2 to 1.0 | 0.158 |
| GCSI<sup>b</sup> | 2.5 (1.0) | 2.4 (1.1) | 0.1 (1.0) | -0.2 to 0.5 | 0.432 |
| Upper Abdominal Pain Score | 2.8 (1.4) | 2.6 (1.7) | 0.2 (1.6) | -0.4 to 0.8 | 0.473 |
| Lower Abdominal Pain Score | 2.5 (1.5) | 2.3 (1.5) | 0.2 (1.5) | -0.3 to 0.8 | 0.360 |
| Heartburn Score | 1.7 (1.4) | 1.4 (1.2) | 0.4 (1.3) | -0.1 to 0.8 | 0.124 |
| **MPQ PRI** |  |  |  |  |  |
| MPQ Sensory | 12 (8) | 11 (8) | 1 (8) | -2 to 4 | 0.394 |
| MPQ Affective | 4.1 (3.6) | 4.3 (3.7) | -0.2 (3.7) | -1.5 to 1.1 | 0.742 |
| **PFDI-20** |  |  |  |  |  |
| POPDI-6 | 27 (23) | 20 (19) | 8 (21) | 0 to 15 | 0.048 |
| CRADI | 31 (23) | 24 (21) | 7 (22) | -1 to 15 | 0.067 |
| UDI | 29 (28) | 18 (22) | 10 (24) | 1 to 19 | 0.025 |
| **PAC-SYM** |  |  |  |  |  |
| Abdominal Score | 2.1 (1.0) | 1.8 (1.0) | 0.3 (1.0) | -0.1 to 0.6 | 0.099 |
| Rectal Score | 0.9 (0.9) | 0.9 (0.9) | 0.0 (0.9) | -0.3 to 0.4 | 0.775 |
| Stool Score | 1.9 (1.1) | 1.5 (1.1) | 0.3 (1.1) | -0.1 to 0.7 | 0.096 |
| **COMPASS-31** |  |  |  |  |  |
| Orthostatic Intolerance Score | 21 (14) | 17 (12) | 4 (13) | 0 to 9 | 0.079 |
| Vasomotor Score | 1.8 (1.8) | 1.0 (1.6) | 0.8 (1.7) | 0.2 to 1.4 | 0.012 |
| Secretomotor Score | 5.3 (3.6) | 4.3 (3.7) | 1.0 (3.7) | -0.3 to 2.3 | 0.141 |
| Gastrointestinal Score | 11.9 (3.8) | 10.6 (4.4) | 1.3 (4.2) | -0.3 to 2.8 | 0.102 |
| Bladder Score | 2.1 (2.1) | 1.3 (1.7) | 0.8 (1.9) | 0.1 to 1.5 | 0.023 |
| Pupillomotor Score | 2.4 (1.4) | 1.8 (1.4) | 0.6 (1.4) | 0.1 to 1.1 | 0.028 |

\*Mean (SD).
<sup>a</sup> Two-sample *t* test.
<sup>b</sup> The GCSI is derived from the three preceding scores
PAGI-SYM = patient assessment of upper gastrointestinal symptom severity index; GCSI = Gastroparesis Cardinal Symptom Index; MPQ PRI = McGill Pain Questionnaire Pain Rating Indices; PFDI = Pelvic Floor Distress Inventory; POPDI = Pelvic Organ Prolapse Distress Inventory; CRADI = Colorectal-Anal Distress Inventory; UDI = Urinary Distress Inventory; PAC-SYM = Patient Assessment of Constipation-Symptoms; COMPASS = Composite Autonomic Symptom Score.

**Supplemental Table 2.** Comparison of symptom subscales between patients with and without joint hypermobility/hypermobility spectrum disorder (JH/HSD)*

Terms in bold indicate the global test scores (displayed in Table 6)
|  | **JH/HSD**<br>**(N = 67)** | **Not JH/HSD**<br>**(N = 66)** | **$\Delta$** | **95% CI** | **P<sup>a</sup>** |
| --- | --- | --- | --- | --- | --- |
| **PAGI-SYM** |  |  |  |  |  |
| Nausea Score | 1.5 (1.1) | 1.4 (1.4) | 0.1 (1.3) | -0.3 to 0.5 | 0.611 |
| Fullness Score | 3.0 (1.3) | 2.6 (1.4) | 0.4 (1.3) | -0.1 to 0.8 | 0.110 |
| Bloating Score | 3.2 (1.5) | 2.8 (1.6) | 0.5 (1.6) | -0.1 to 1.0 | 0.101 |
| GCSI<sup>b</sup> | 2.6 (1.0) | 2.3 (1.1) | 0.3 (1.0) | 0.0 to 0.7 | 0.091 |
| Upper Abdominal Pain Score | 2.9 (1.5) | 2.4 (1.7) | 0.6 (1.6) | 0.0 to 1.1 | 0.043 |
| Lower Abdominal Pain Score | 2.7 (1.5) | 2.1 (1.5) | 0.6 (1.5) | 0.1 to 1.1 | 0.020 |
| Heartburn Score | 1.8 (1.3) | 1.2 (1.3) | 0.6 (1.3) | 0.1 to 1.0 | 0.009 |
| **MPQ-PRI** |  |  |  |  |  |
| MPQ Sensory | 13 (8) | 9 (8) | 4 (8) | 1 to 6 | 0.010 |
| MPQ Affective | 4.5 (3.4) | 4.0 (3.9) | 0.5 (3.7) | -0.8 to 1.7 | 0.438 |
| **PFDI-20** |  |  |  |  |  |
| POPDI | 28 (22) | 17 (19) | 11 (20) | 4 to 18 | 0.003 |
| CRADI | 32 (22) | 21 (20) | 11 (21) | 4 to 18 | 0.003 |
| UDI | 25 (26) | 19 (24) | 6 (25) | -2 to 15 | 0.133 |
| **PAC-SYM** |  |  |  |  |  |
| Abdominal Score | 2.1 (1.0) | 1.7 (0.9) | 0.5 (1.0) | 0.1 to 0.8 | 0.005 |
| Rectal Score | 1.1 (0.9) | 0.8 (0.8) | 0.3 (0.9) | 0.0 to 0.6 | 0.052 |
| Stool Score | 2.0 (1.1) | 1.4 (1.0) | 0.6 (1.0) | 0.2 to 1.0 | 0.001 |
| **COMPASS** |  |  |  |  |  |
| Orthostatic Intolerance Score | 23 (12) | 13 (11) | 10 (12) | 6 to 14 | <0.001 |
| Vasomotor Score | 1.8 (1.8) | 0.8 (1.5) | 1.0 (1.7) | 0.4 to 1.5 | 0.001 |
| Secretomotor Score | 5.3 (3.5) | 4.0 (3.8) | 1.3 (3.6) | 0.0 to 2.5 | 0.049 |
| Gastrointestinal Score | 12.1 (3.8) | 10.1 (4.4) | 2.0 (4.1) | 0.6 to 3.4 | 0.006 |
| Bladder Score | 1.9 (2.1) | 1.2 (1.6) | 0.7 (1.9) | 0.1 to 1.4 | 0.027 |
| Pupillomotor Score | 2.5 (1.4) | 1.5 (1.3) | 1.0 (1.4) | 0.5 to 1.4 | <0.001 |

\*Mean (SD).
<sup>a</sup>Two-sample *t* test.
<sup>b</sup> The GCSI is derived from the three preceding scores
PAGI-SYM = patient assessment of upper gastrointestinal symptom severity index; GCSI = Gastroparesis Cardinal Symptom Index; MPQ PRI = McGill Pain Questionnaire Pain Rating Indices; PFDI = Pelvic Floor Distress Inventory; POPDI = Pelvic Organ Prolapse Distress Inventory; CRADI = Colorectal-Anal Distress Inventory; UDI = Urinary Distress Inventory; PAC-SYM = Patient Assessment of Constipation-Symptoms; COMPASS = Composite Autonomic Symptom Score.

**Supplemental Table 3.**
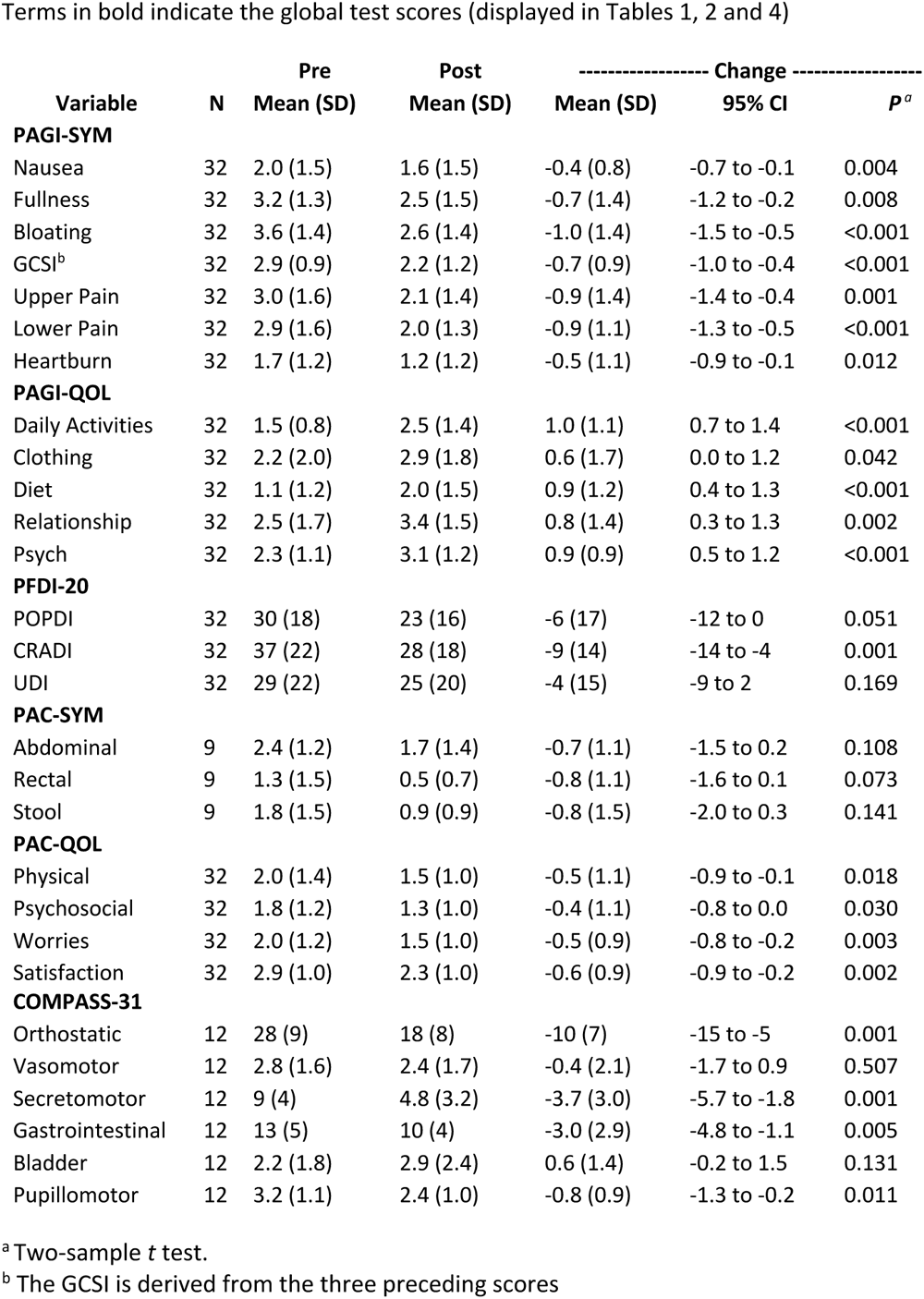
Change in subscale score following IVIG treatment.

### APPENDIX: Methods

#### QUESTIONNAIRES

##### PAGI-SYM and GCSI

The 20-item patient assessment of upper gastrointestinal symptom severity index (PAGI-SYM) has six subscales: heartburn/regurgitation, fullness/early satiety, nausea/vomiting, bloating, upper abdominal pain, and lower abdominal pain measured over the preceding two weeks and scored on a scale of severity (from 0 to 5).^1^ The Gastroparesis Cardinal Symptom Index (GCSI), in patients that utilizes three symptom clusters within the PAGI-SYM (nausea/vomiting/retching, post-prandial fullness/satiety, and bloating/stomach distention).^2^

##### PAGI-QOL

The Patient Assessment of Upper Gastrointestinal Disorders-Quality of Life (PAGI-QOL) is a 30-item instrument that has been validated as a reliable and sensitive measure of quality of life in patients suffering from dyspepsia, GERD or gastroparesis in the prior two weeks.^3^ Higher scores indicate better health-related quality of life.

##### PAC-SYM

The Patient Assessment of Constipation Symptoms (PAC-SYM) questionnaire is a 12-item questionnaire is divided into three symptom subscales: abdominal (four items); rectal (three items); and stool (five items). Items are scored on 5-point Likert scales, with scores ranging from 0 to 4 (0 = ‘symptom absent’, 1 = ‘mild’, 2 = ‘moderate’, 3 = ‘severe’ and 4 = ‘very severe’).^4^

##### PAC-QOL

The PAC-QOL is a self-administered questionnaire for the assessment constipation related impact on quality of life. The overall and all subscale scores range from 0 to 4, with lower scores indicating better health-related quality of life.^5^

##### PFDI-20

The Pelvic Floor Distress Inventory Questionnaire-20 (PFDI-20) is a patient reported outcome measure for assessing the distress associated with pelvic floor disorders. It consists of 3 subscales, which include the Urinary Distress Inventory-6 (UDI-6), Pelvic Organ Prolapse Distress Inventory-6 (POPDI-6), and the Colorectal-Anal Distress Inventory-8 (CRADI-8).^6^

##### Eckardt Dysphagia Symptom Score

This is a self-reported scale measuring weight loss in kilograms, chest pain, regurgitation, and dysphagia. Each of these 4 items is graded on a score of 0 to 3.^7^

##### COMPASS-31

The Composite Autonomic Symptom Score (COMPASS) 31 is a self-assessment instrument for assessing symptoms related to autonomic nervous dysfunction and consists of six domains: orthostatic intolerance (four questions); vasomotor (three questions); secretomotor (four questions); gastrointestinal (12 questions); bladder (three questions); and pupillomotor (five questions)^.8^

5-point questionnaire (5-PQ) score for joint hypermobility/hypermobility spectrum disorder This is a self-reported instrument in the form of a five-part questionnaire (5PQ) that includes five aspects of past or present hypermobility, with a cut-off level of two positive answers to any of the five questions.9 It has been validated for use in patient cohorts in the clinic10 as well as population-based studies.11 It advantages over the Beighton score is that it does not require a physical examination, checks for previous agility, and therefore provides a more generalized assessment of hypermobility, rather than focusing exclusively on 5 joints. False negatives are expected to be less with this approach and its use has been accepted as an alternative way to identify patients with hypermobility spectrum disorders (HSD).^12^

##### Short form McGill Pain Questionnaire (SF-MPQ)

The SF-MPQ consists of 15 representative words from the sensory (n = 11, consisting of descriptors of the sensory qualities of the pain experience) and affective (n = 4, consisting of descriptors that describe the emotional/affective or autonomic qualities) of the standard, Long-Form MPQ (LF-MPQ). Each descriptor is ranked by the patient on an intensity scale of 0 = “none,” 1 = “mild,” 2 = “moderate,” 3 = “severe. The PRI (T) or Total Pain Intensity is obtained by the sum of the sensory and affective PRIs. The PPI (present pain intensity) and a visual analog scale (VAS) are also included.^13^

##### PROMIS® (Patient-Reported Outcomes Measurement Information System) Scales (https://www.healthmeasures.net/)

For all PROMIS scales in this study, the final score is represented by the T-score, a standardized score with a mean of 50 and a standard deviation (SD) of 10. This means that a score of 50 represents the average of the general population (and that 10 represents the standard deviation). A higher PROMIS T-score represents more of the concept being measured. For negatively-worded concepts like Anxiety, a T-score of 60 is one SD worse than average. By comparison, an Anxiety T-score of 40 is one SD better than average. However, for positively-worded concepts like Physical Function-Mobility, a T-score of 60 is one SD better than average while a T-score of 40 is one SD worse than average.

The PROMIS Gastrointestinal Belly Pain Scale (PROMIS Scale v1.0 - GI Belly Pain 5a) assesses the severity of belly pain over the past 7 days. It is useful for pain that varies in location, intensity, and quality as it is not region or disease specific. Severity is a composite of several factors including intensity, frequency, bothersomeness, nature, predictability and number of regions affected^.14^

The PROMIS Gastrointestinal Bowel Incontinence Scale (PROMIS Scale v1.0 - Gastrointestinal Bowel Incontinence 4a) assesses the frequency of bowel incontinence, soiling, and gas incontinence (i.e. stool leakage while passing gas) over the past 7 days.14

The PROMIS Gastrointestinal Diarrhea Scale (PROMIS Scale v1.0 - Gastrointestinal Diarrhea 6a) contains items focused on capturing the frequency and form of the stool and its bothersomeness, impact, controllability, and predictability. during the past 7 days.^14^

The PROMIS Gastrointestinal Gas and Bloating Scale (PROMIS Scale v1.1 - GI Gas and Bloating 13a) assesses the frequency and intensity/severity of bloating (i.e., feeling pressure or fullness), bloating appearance (i.e., belly swollen or larger than usual size), flatulence (i.e., passing gas), and abdominal sounds (i.e., gurgling or rumbling). The scale also assesses the degree of bother and interference with daily activities resulting from bloating and swelling.^14^

PROMIS Nociceptive and Neuropathic Scores (PROMIS Scale v2.0 - Nociceptive Pain Quality 5a and PROMIS Scale v2.0 - Neuropathic Pain Quality 5a)

Both scales consist of 5 items rated on a 5-point Likert-type from “not at all” to “very much.” Responses are based on the participant’s symptoms over the last 7 days. Descriptors of neuropathic pain include “pins and needles,” “tingly,” “stinging,” “electrical,” and “numb.” Descriptors of nociceptive pain, include “sore,” “tender,” “achy,” “deep,” and “steady.^15^

PROMIS physical function, anxiety, depression, fatigue, sleep disturbance, satisfaction with participation in social roles, pain interference and a 0-10 pain intensity scale are part of the 7 domains in the PROMIS-29 Profile v2.0.^16^

##### Headache Screeing Questionnaire for Migraine and Tension Type Headache (TTH)

The HSQ-DV is a short 10-item screening tool that can be used for recognition of probable migraine (sensitivity 0.89) and probable TTH (sensitivity 0.92). A score of > 6 indicates “probable” migraine or TTH and a score of 8 indicates definite migraine or TTH.17

##### Overall Treatment Effect (OTE)

For assessing the global response to IVIG treatment, patients were asked the following question “Since your last infusion, has there been any change in activity, limitations, symptoms, emotions, or overall quality of life related to your illness?”^18^

No change= 0

A very great deal worse -7

A great deal worse -6

A good deal worse -5

Moderately worse -4

Somewhat better -3

A little worse -2

Almost the same, hardly any worse at all -1

Almost the same, hardly any better at all 1

A little better 2

Somewhat better 3

Moderately better 4

A good deal better 5

A great deal better 6

A very great deal better 7

#### DIAGNOSTIC TESTING

##### Whole gut scintigraphy

This was single gastrointestinal transit study including esophageal transit, liquid and solid gastric emptying, and small-and large bowel transit, using 111In-diethylenetriaminepentaacetic acid (DTPA), as previously described.^19^

##### Skin biopsy

Skin biopsies were performed at standard sites (proximal thigh, distal thigh, and distal leg) and immunohistochemical techniques were used to identify intraepidermal nerve fiber density and sudomotor (sweat gland) innervation were identified using previously published normative data established by the laboratory.^20–22^

##### Tilt Table Test

The Tilt Table Test was performed on pediatric and adult patients who, have a clinical presentation compatible with dysautonomia. Patients were less than 400 pounds in weight, were at least 4 hours without food, and, if a female of childbearing potential, a negative HCG urine or blood test was negative. All patients have peripheral IV access, and placement of a semi-automatic non-invasive blood pressure system, pulse oximeter, and 12 lead ECG leads. The tilt test started with a 15-minute supine baseline period consisting of recording vital signs and ECG strips every 5 minutes. The average of pulse rate and blood pressure was used as the baseline hemodynamics. Stage 1 of the tilt test starts immediately at the end of the supine baseline period when the table is positioned at a 60-80 degree tilt. Observations of vital signs are taken at immediate upright tilt, and 2 minutes, 5 minutes and then every 5 minutes after that until the last at 45 minutes. During each observation period the nurse inquired as to the presence of symptoms or comments from the patient. The nurse watched the continuous ECG tracing and obtained additional sampling of the vital signs at any time during the 5-minute observation periods if thought necessary.

Hemodynamic criteria for postural orthostatic tachycardia syndrome (POTS) were an increase in heart rate during the first 10 minutes of upright tilt of 30 bpm of tilt for individuals of 20 years of age or older. For those younger than 20 years of age the rate increment is 40 bpm or more. An ancillary criterion for POTS is a heart rate of 120 bpm or more in the first 10 minutes of tilt at any age. The hemodynamic criteria for neurally mediated hypotension were a drop in systolic blood pressure of 20 mmHg or more associated with symptoms. If the patient did not have NMH during stage 1 he/she is returned to the supine position and stage 2 of the tilt test is started with a 10-minute supine infusion of isoproterenol (2 mcg/min) with a new baseline established prior to repeating upright tilt. The duration of stage 2 was 15 minutes if not ended prior to that time with NMH. After the tilt test the patient received IV fluids (usually 1 liter of normal saline) in the post procedure care unit and met with the physician to discuss the test results and counseled as to what that might mean.

## Notes

### Competing Interest Statement

The authors have declared no competing interest.

### Clinical Trial

NCT04859829

### Funding Statement

Funded in part by the Amos Food Body and Mind Center, Johns Hopkins University School of Medicine

### Author Declarations

The entire study was approved by the Institutional Review Board of the Johns Hopkins University School of Medicine

### Summary of Updates

Removed a duplicate entry in Table 1

